# Beyond the Target Product Profile: Design Requirements for Point-of-Need HCV RNA Testing in Harm Reduction Settings

**DOI:** 10.64898/2026.08.14.26357799

**Authors:** Samantha Mata-Robles, Khalid Khalaf, Jillian Kelley, Akansha Chauhan, Lara Balian, Jacqueline C Linnes, Natalia M Rodriguez

## Abstract

Point-of-care Hepatitis C Virus (HCV) RNA assays reduce diagnostic turnaround time but depend on benchtop instrumentation and continuous electricity, limiting their deployment in the harm reduction and community settings where confirmatory testing is most needed, as people who use drugs (PWUD) carry a disproportionate share of the HCV burden in the United States. This is a systemic problem in diagnostic development, where decision-making and design requirements overlook point-of-use stakeholders. Closing the gap requires integrating real-world constraints throughout design rather than validating against user needs once a product already exists. Here, we apply a human-centered design (HCD) approach to inform rigorous, stakeholder-derived design requirements, implementation considerations, and early value proposition for a novel point-of-need HCV RNA test intended for deployment in harm reduction and community settings in Indiana. To determine design specifications grounded in real-world context, our objectives were (1) identifying and characterizing context-specific experiences and barriers to HCV testing among higher-risk populations; (2) assessing the perceived benefits and acceptability of the proposed test within real-world settings across direct and indirect user groups; and (3) translating the user needs and contextual constraints into design requirements and implementation considerations that support the test’s clinical, operational, and user-centered value. We conducted 18 semi-structured interviews with frontline staff and HCV testing/treatment pipeline experts (n=11) and people who get tested (n=7) across harm reduction organizations, syringe service programs, and community testing settings, analyzed using Rapid Qualitative Analysis guided by the PARRQA framework. Stakeholders responded positively to a single-encounter point-of-need RNA test, and implementation considerations, including funding restrictions, staffing structures, and diverse deployment settings, directly shaped design requirements spanning turnaround time, sample type and volume, portability, result output, target operator, and ease of use. Benchmarking these stakeholder-derived specifications against the FIND Dx HCV target product profile (TPP) showed that stakeholder input confirmed, modified, or extended several TPP criteria and introduced requirements the TPP does not address. Together, these objectives constitute an upstream, evidence-driven design process that translates contextual and stakeholder knowledge into actionable engineering requirements, highlighting the need for diverse stakeholder engagement at all stages of the design process for closing the translation gap between laboratory-validated diagnostic tools and effective point-of-need deployment.

## Introduction

The Hepatitis C Virus (HCV) is the leading cause of liver disease in the US, making it a major public health problem (1,2). This bloodborne pathogen causes inflammation of the liver that, without timely diagnosis and treatment, leads to irreversible outcomes including cirrhosis, hepatocellular carcinoma, and in the worst cases, death (3–5). However, clinically apparent hepatic damage typically spans two to three decades, offering a critical window for timely diagnosis and treatment initiation (6,7). Now, direct-acting antiviral therapies achieve cure rates exceeding 95%, meaning that every undetected infection represents preventable harm (8).

HCV infection affects an estimated 2.4 million adults in the United States, though this figure likely underestimates the true burden given known gaps in surveillance coverage among incarcerated and unhoused populations (9–11). Despite the availability of diagnostic and treatment options, up to 75% of individuals living with HCV remain unaware of their infection (12). However, this burden is not uniformly distributed. National surveillance data identified injection drug use as the primary driver of HCV transmission in the United States. Indiana exemplifies this pattern; the state ranked 11th in the nation for rates of reported acute HCV infection (13), with approximately 68% of reported cases indicating a history of injection drug use (14). This disparity can be attributed to limited access to HCV testing and linkage to care, highlighting the need to facilitate full diagnosis and treatment initiation to close this gap (15).

The standard diagnostic pathway recommended by the CDC follows a two-step testing protocol, initiated by screening with a Food and Drug Administration (FDA) - approved antibody test, if reactive it is followed by a nucleic acid amplification test (NAT) to detect HCV RNA (16). There is documentation that demonstrates how this method limits effective diagnosis and linkage to care. According to the Chronic Hepatitis Cohort Study, only 62% of patients complete the full CDC-recommended diagnostic pathway (17). The Veterans Health Administration reported a similarly low completion rate of 64%, compared to 98% in facilities where automatic HCV RNA testing was performed (17), demonstrating that the protocol gap is not a patient behavior problem, but a systems design problem.

Recent advances in molecular diagnostics have led to the development of near-patient RNA testing platforms that reduce turnaround time. The first FDA-authorized point-of-care HCV RNA test, the Xpert® HCV Viral Load Fingerstick assay (Cepheid), enables detection of active infection from a capillary blood sample within approximately one hour (18,19). Non–FDA–approved HCV RNA near-patient systems, including the Genedrive® HCV RNA test and Truenat® HCV platform, have demonstrated feasibility for decentralized testing and have received regulatory approval or prequalification for use outside the United States (20–24). These tests require benchtop instrumentation to be purchased or leased, a minimal laboratory setup with continuous electricity for proper performance.

While these point-of-care tests show great promise and are viable options to be implemented in clinical settings, their dependence on benchtop instrumentation, a minimal laboratory setup with continuous electricity for its proper performance, keeps them from benefiting the populations that need them the most, especially people who use drugs (PWUD) (24,25). This opens a gap for the development of a point-of-need test that enables diagnosis outside of hospital or clinical environments. As established above, this population carries a disproportionate share of the HCV burden. Housing instability, criminalization, and competing survival priorities such as securing food, shelter, and safety from violence make it difficult for this population to return for a second clinical visit or travel to a centralized laboratory (26,27). Harm reduction organizations are built around meeting people where they are, delivering services without requiring stable housing, insurance, or a scheduled follow-up (28–30). This would allow for the test to be deployed in syringe service programs (SSPs) or harm reduction organizations where single-encounter results are most needed (25,26).

To address this gap, we are working towards developing a molecular diagnostic assay paired with a lateral flow test, integrated in a handheld device for the point-of-need detection of HCV RNA in community settings. To guide the design and implementation, the existing target product profile (TPP) defines minimal and optimal characteristics to be met, such as intended use and target populations, as well as performance and operational characteristics (31,32). These requirements developed by FIND Dx involved diverse stakeholders, including technical experts, international and national organizations, as well as implementers. While this document provides useful information for broadly applicable test development, it lacks the direct engagement of harm reduction staff, community health workers (CHWs), or PWUD in specific US community settings, which leaves a gap for what our target population actually needs, values, and will accept in practice. This is reflected in how, despite the FIND Target Product Profile for HCV decentralized testing existing since 2015 (31), and molecular point-of-care assays such as the Genedrive HCV meeting its performance specifications (21,22,24), HCV diagnosis and treatment rates among PWUD remain disproportionately low (15,33). While the opioid epidemic has contributed to this continuous increased rate of new HCV cases since 2013 (33,34), this also suggests the TPP does not capture all of the necessary information for successful real-world adoption at the point of need to close this gap.

This is a systemic problem in diagnostic development, where decision-making and design requirements overlook point-of-use stakeholders. Closing the gap requires integrating real-world constraints throughout design rather than validating against user needs after a product already exists. This study applies a human-centered design (HCD) approach to inform rigorous, stakeholder-derived design requirements, implementation considerations, and early value proposition for a novel point-of-need HCV RNA test intended for deployment in harm reduction and community settings in Indiana. To determine design specifications grounded in real-world context, our objectives were (1) identifying and characterizing context-specific experiences and barriers to HCV testing among higher-risk populations; (2) assessing the perceived benefits and acceptability of the proposed test within real-world settings across direct and indirect user groups; and (3) translating the user needs and contextual constraints into design requirements and implementation considerations that support the test’s clinical, operational, and user-centered value. Together, these objectives constitute an upstream, evidence-driven design process that translates contextual and stakeholder knowledge into actionable engineering requirements.

## Methodology

### Study Design

HCD consists of three different phases, Inspiration where we first work to understand who we are designing for; Ideation where ideas to potential solutions are generated and prototyped; and finally, Implementation, where the idea gets piloted, turned into a reality, and translated into a fully feasible and viable solution (35–37). To develop stakeholder-grounded design specifications, we employed the inspiration and ideation phases consisting of contextual investigation and qualitative stakeholder interviews analyzed using rapid qualitative analysis. HCD was selected for its capacity to engage direct and indirect stakeholders whose constraints are not captured in top-down approaches such as those implemented to develop the FIND Dx TPP (31).

### Immersion in the Context

To better inform our interview questions, we immersed ourselves in different community settings to observe and better understand the different audiences our solution would affect. We spent multiple days shadowing the testing process for HCV in two counties in Indiana, and several testing settings, including harm reduction organizations, a housing organization, recovery events and organizations, and the organization’s offices. Field notes were taken and the observations and lessons learned from this experience informed the interview guide development for our semi-structured interviews.

### Sampling and participants

For the inspiration phase of the HCD approach, frontline staff (FS) including testers and HCV testing and treatment pipeline experts; ranging from community health workers that offer testing for PWUD, case managers, community organization leaders, subject matter experts, and clinicians; were recruited through snowball sampling. Recruitment flyers and information were sent to people within our research group’s network and publicly available emails from organizations that work in the Hepatitis C pipeline amongst higher-risk populations in Indiana. These participants were prioritized for the study, which required working with HCV diagnosis testing or integration to care in Indiana; however, they were not eligible if they did so outside of Indiana. People who get tested (PWGT) for HCV were recruited through community partner organizations in Indiana using a script to ensure no coercion and voluntary participation. These participants were eligible to participate in the study if they were over 18 years old, used harm reduction services in Indiana, and had heard of HIV/HCV testing. As an incentive for participation, all candidates were offered a $50 Visa gift card.

### Data collection

The semi-structured interviews with the frontline staff and HCV testing and treatment pipeline experts were conducted through Zoom; whereas the ones conducted with people who get tested were done in-person at partner organizations in separate private rooms. All interviews were conducted only in the presence of the participant and researchers, and they were voice-recorded and transcribed using Otter.ai.

### Analysis

Rapid Qualitative Analysis (RQA) (38) was selected as the analytic method for the ideation phase of the HCD approach given the study’s narrow scope, focused research questions, highly structured data collection processes, and targeted semi-structured interview protocols. The Planning for and Assessing Rigor in Rapid Qualitative Analysis (PARRQA) framework was developed by qualitative methods experts to support rigor and validity in projects using RQA, including study design, conduct, write-up, and review (39). This framework is broken down into five phases, (1) rigorous design where the rationale for the study, methodology and approach are established, (2) semi-structured data collection where the materials to be used are developed, tested, and deployed, (3) RQA where the summary templates for the data analysis are developed and calibrated, (4) RQA matrices development for data centralization, (5) RQA data synthesis.

RQA was conducted separately for each stakeholder group, one for the frontline staff, and one for people who get tested, resulting in two independent analytic processes. The semi-structured interview guide questions were framed and mapped to key areas of interest including HCV testing experiences, challenges faced, barriers, perceived acceptability, design preferences, implementation considerations, and needs; leading to the domains to be used for the summary template and our analysis. The summaries were completed by listening to the audio files and linking the relevant information to the corresponding domains. For the data centralization and synthesis, a matrix was constructed per stakeholder group in Microsoft Excel with interview IDs as rows and thematic domains as columns. Each domain corresponded to a designated area of inquiry from the interview guide. A pair of researchers was assigned to each stakeholder matrix (38,39). Each pair independently reviewed and summarized interview transcripts, extracting responses relevant to each domain, and reached consensus on the final summaries through discussion. To establish rigor and cross-matrix consistency, both completed matrices and their corresponding analyses were reviewed by a third researcher on the team, who provided elaboration or expansion where warranted. The resulting matrices enabled systematic comparison of responses across participants within each stakeholder group, as well as identification of patterns and divergences between stakeholder perspectives. The data reporting in this manuscript was done following the COREQ (COnsolidated criteria for REporting Qualitative research) checklist (40).

### Researcher Positionality

The research team is composed entirely of academic researchers in biomedical engineering and public health at Purdue University, with no team member reporting personal or lived proximity to harm reduction services or injection drug use. The team’s relationship with community partners was established through pre-existing professional networks in HIV-linked organizations, through which the team was introduced to a frontline harm reduction tester in Indiana, who in turn connected the team to additional organizations working in this space. Prior to data collection, the lead researcher (SMR) conducted an immersion period with frontline staff, shadowing and holding informal conversations at harm reduction organizations and syringe service programs to build familiarity with the operational context and establish rapport. Partnering organizations agreed to participate on the basis of a shared commitment to the populations they serve. RQA was conducted independently for each stakeholder group by two analyst pairs (SMR and JK for one group, SMR and AC for the other), with KK serving as a third reviewer across both, and the interview guide was reviewed prior to deployment by a team member trained in RQA methodology (LB), which provided an additional check on content coverage before interviews began. The team’s disciplinary orientation toward diagnostic device design shaped the interview guide and analytic domains toward operational and technical design requirements. This focus was deliberate as a substantial existing literature documents lived experience narratives among people who use drugs and people affected by HCV. This study’s aim was to characterize how participants’ situations and contexts shaped their testing experiences, generating actionable design and implementation specifications for a point-of-need diagnostic.

### Ethical considerations

IRB of Purdue University gave ethical approval for this work.

Institutional approval for this study was obtained through Purdue University (Frontline staff IRB-2024-521 and People who get tested IRB-2024-1763). To protect the participants, their capacity for consent was assessed by asking them five comprehensive questions about the study; if answered correctly, the participants were eligible to be part of the study. To mitigate risk of breach of privacy names were not recorded during the interviews; consent was obtained prior to beginning the recording and once again once the recording had started. All data was de-identified before storage; and all handling was done by trained personnel and stored in a secure password protected folder.

## Results

Results are presented in the following order: participant characteristics, contextual investigation, perceived acceptability and value proposition, implementation considerations, design considerations, and lastly translating the findings to design specifications. This order was intentional as the contextual investigation establishes the environment in which testing currently occurs, perceived acceptability establishes whether the proposed solution is needed and how stakeholders initially responded to it, and implementation considerations establish the real-world constraints the device must operate within. Each of these sections informs the design considerations presented last, which are then translated into the design specifications in Table 3.

### Participant Characteristics

A total of 11 semi-structured interviews were conducted with people who work in the HCV diagnosis, testing, and/or integration to care; including health/care managers or coordinators (n=3), harm reduction and preventive services managers (n=2), health educators (n=4), one executive director, and a public health expert. Participants served populations across multiple rural and urban Indiana counties, including people who inject drugs, unhoused individuals, formerly incarcerated individuals, and LGBTQ+ community members including men who have sex with men, and sex workers.

A total of seven interviews were conducted with people who get tested (PWGT) in Indiana (n=7) (Fig. 1). Four participants identified as male and three as female. Five participants reported their race as White/Caucasian, while the remaining two did not answer. Three participants were employed at the time of the interview. Five participants reported being housed, two of whom had recently obtained housing from a recovery organization. All participants reported having insurance; two of them did not specify their provider, two reported having Medicaid, two Healthy Indiana Plan, and one Medicare.

**Figure 1.**
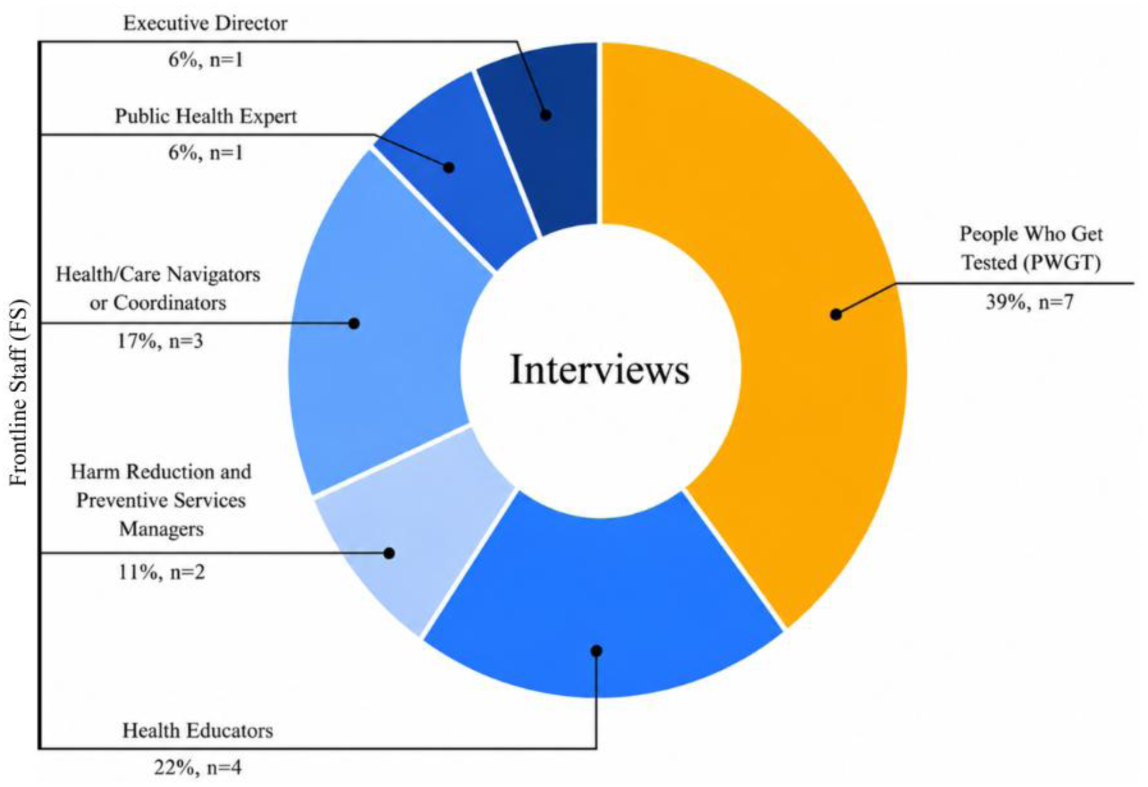
Composition of interview participants by stakeholder group and role (N=18). Frontline staff (n=11, blue) included Health Educators, Health/Care Navigators or Coordinators, Harm Reduction and Preventive Services Managers, Public Health Experts, and Executive Directors. People Who Get Tested (39%, n=7, yellow) were interviewed as a separate stakeholder group, consistent with the two independent RQA analytic processes described in the Methods.

### Contextual Investigation

Organizations across Indiana work to deliver point-of-care testing to help diagnose and treat those more at need using an FDA approved rapid antibody test, the Oraquick HCV Rapid Antibody Test. The organization’s HCV-related programs are funded by state-level grants, meaning that the expenses are bound by their guidelines; limiting what can be done with the funds. In most cases, the grants allow organizations to purchase rapid antibody tests; however, not all other important expenses are covered. Supplementary programs to mitigate competing priorities, such as transportation with gas money, and food pantry, fall outside grant stipulations; impacting what organizations can offer beyond the test itself, limiting the follow-up support available once a reactive result is obtained. The point-of-care test is offered across diverse settings, including harm reduction organizations, community events, and street outreach. Testing organizations collaborate extensively with community partners, including transitional housing programs, warming centers, churches, libraries, food pantries, jails, and SSPs, to reach clients who would not otherwise access services.

The testing process takes 15-30 minutes, and it involves multiple steps including consent, risk behavior assessment, running the antibody test, and educational content and sharing of health information while waiting for test results. However, this test is only the first step in a diagnostic pathway that the current infrastructure isn’t equipped to complete. If the test comes back reactive, the patient information is then shared with a case manager who is usually not on site. Case managers must later try to reach out to the patient to schedule the confirmatory RNA test at a separate facility. The stakeholders mentioned that many of their patients are unhoused, have limited phone access, and are unemployed or work minimum-wage jobs; which was also reflected when we talked to the people who get tested; circumstances that make outreach and follow-up extremely challenging. The full diagnostic pathway, from reactive antibody result to a confirmatory molecular test, normally spans a couple of weeks or longer, further increasing the probability of disengagement at every step. While patient loss occurred at every stage, the highest was between a reactive antibody result and the completion of confirmatory molecular testing. (Fig. 2). Testing organizations recognize this failure point and develop interventions to support patients in remaining engaged in the testing pathway and treatment, including refillable prepaid debit cards for transportation and food. However, as discussed earlier, these are limited to grant stipulations.

**Figure 2.**
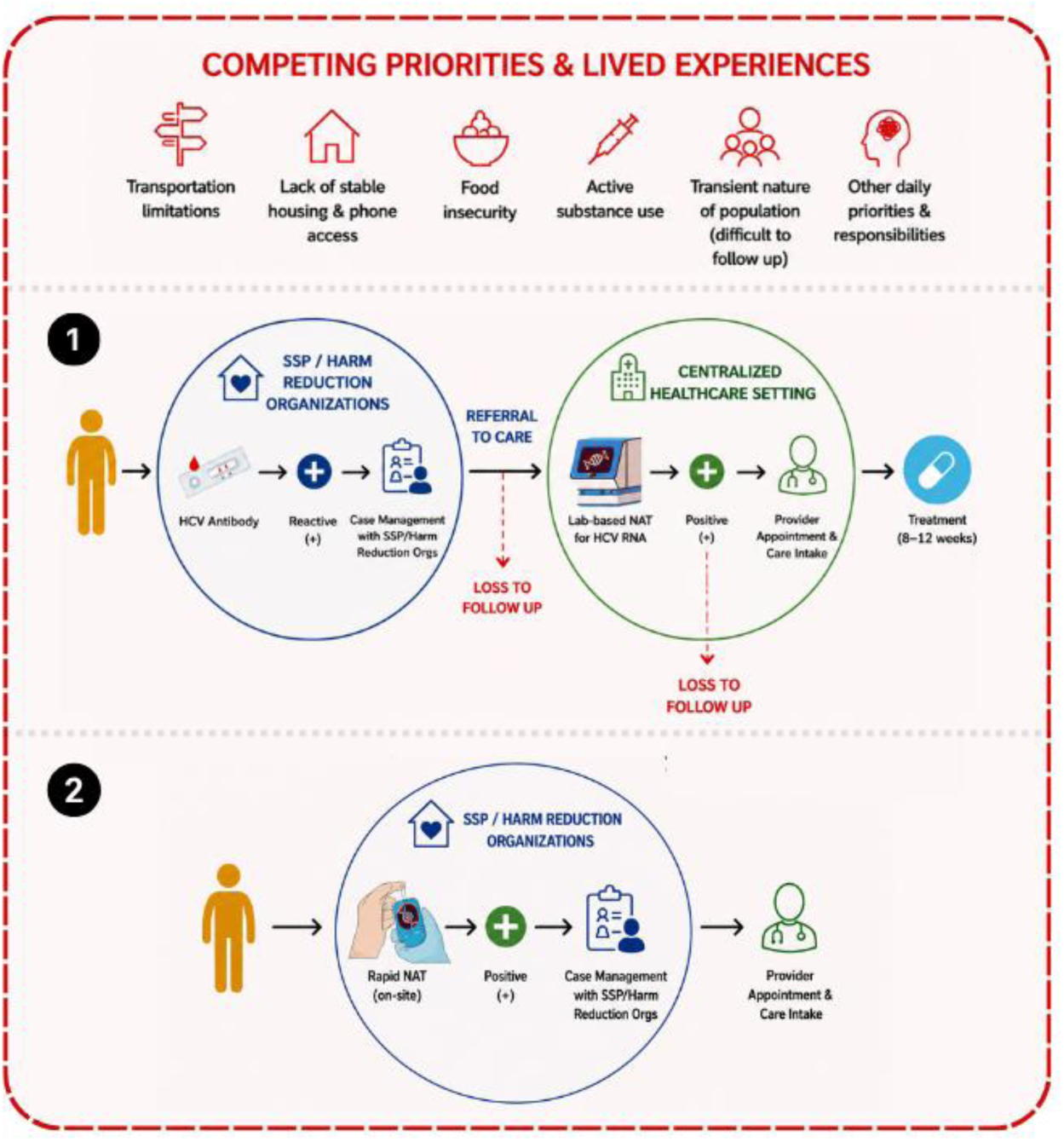
Current and proposed HCV diagnostic pathways in point-of-need testing settings. (1) Current pathway: a person tested at an SSP or harm reduction organization receives an HCV antibody rapid diagnostic test on-site. A reactive result triggers case management and referral to a centralized healthcare setting for a second test, the laboratory-based confirmatory RNA testing; this is the first point of loss to follow-up. A positive RNA result then requires a separate provider appointment and care intake before an 8–12 week treatment course can begin, a second point of loss to follow-up. (2) Proposed pathway: a point-of-need rapid RNA test performed on-site at the SSP or harm reduction organization returns a same-encounter result, allowing case management and provider appointment and care intake to proceed directly at the point of testing.

Participants described barriers at multiple levels of the social-ecological model (41,42) throughout the testing and diagnostic pathway. At the individual level, barriers included distrust of the healthcare system, misinformation about HCV risk and transmission, active substance use, and concerns about treatment effects on the body. At the interpersonal level, the normalization of HCV within drug-using social networks reduced the perceived urgency to seek testing, although a single participant reported not being a drug user but living with one and not only being aware of HCV but also being hypervigilant about it. Social isolation from family was common, though two participants described meaningful support from a family member and a peer who had personal treatment experience. Most consistently, harm reduction staff emerged as a primary source of interpersonal encouragement; participants described being motivated to get tested because staff who knew them personally expressed genuine concern for their health. At the community level, geographic barriers were prominent: frontline staff reported driving to secluded rural areas to offer testing, and participants described the distance to confirmatory RNA testing sites, combined with a lack of reliable transportation, as a significant obstacle to completing the diagnostic pathway. Access to treatment while incarcerated was an additional community-level constraint raised by participants. At the institutional level, the multi-step diagnostic process and insurance navigation requirements were identified as barriers to care engagement; for uninsured patients, case managers absorb the burden of insurance coordination, adding pressure to an already constrained follow-up infrastructure. At the policy level, grant restrictions limited what organizations could offer beyond the test itself. While sobriety is no longer required for HCV treatment eligibility, this policy change had not reached many patients, a gap between policy and practice that frontline staff encountered repeatedly in the field. These barriers together make every additional step in the diagnostic pathway a potential point of loss for this population.

### Perceived Acceptability and Value Proposition

When directly addressed, stakeholders across both groups responded positively to the proposed PON HCV RNA test. The primary perceived value was its potential to consolidate the diagnostic process into a single visit; a positive RNA result obtained on-site would enable immediate patient intake, care coordination, and provider linkage without the delays and loss to follow-up associated with laboratory-based confirmatory testing. From the perspective of people who get tested, the most immediate value was receiving a definitive result within a single encounter, participants described the current wait between a reactive antibody result and confirmatory testing as a significant source of anxiety, with one noting that a 60-minute wait would be "amazing" compared to the weeks currently required.

Frontline staff also highlighted that RNA-based detection addresses the inability of the current antibody-only approach to distinguish active infection from past resolved infection, as well as being able to detect infections at earlier stages, which was important for PWGT when seeking testing as a consequence of a known exposure. Both frontline staff and PWGT also noted the potential for the new test to reduce loss to follow-up and to calm post-exposure anxiety in clients awaiting confirmatory results. While stakeholder response was strongly positive, several frontline staff participants raised important questions about the detection window and analytical accuracy of the proposed technology that would require validation before deployment. Additional tests beyond the RNA result, such as genotyping, liver function, and kidney function panels, would still be needed for full clinical workup.

### Implementation Considerations

Implementation considerations have traditionally targeted public health practitioners; however, because these constraints directly shape what the device needs to do, they are bound to affect design decisions, making them extremely relevant for technology developers. The findings presented in Table 1 characterize the operational environment within which a PON HCV RNA test must function across Indiana’s harm reduction and community testing landscape.

**Table 1.** Implementation considerations for point-of-need HCV RNA testing in Indiana’s harm-reduction and community testing landscape.

| Theme | Subtheme | Supporting Evidence |
| --- | --- | --- |
| Access and Equity | Procurement and supply chain through existing channels | Current Oraquick supplied by state via credit system through outreach supply website; Connect2Cure grant funds testing for some organizations. A parallel state-level or grant-funded procurement pathway should be developed for the new test. |
| Access and Equity | Cost should be less than current options | Test procured through grant, while cost is unknown it was mentioned by frontline staff that it should be less than current test cost. Current Medicaid reimbursement rate is \$42.84 per test in Indiana (43) |
| Setting and Deployment | Viable for both clinical/office and community/field settings | 60-min wait is manageable in a clinical setting but requires additional planning in the field. Field testing is essential as organizations cannot reach transient populations without going into the community. Most testing events are 2–4 hours; design must support this workflow. |
| Independent of laboratory | No constant power supply, no cold chain requirements, minimum biohazard waste | No constant power supply, ideally battery operated that lasts full testing event duration (2–4 hours)<br>No access to temperature controlled environments<br>Frontline staff transport biohazard waste with them post-testing events |
| Testing Environment | Co-location with harm reduction and recovery services | Multiple PWGT already visit harm reduction organizations for co-located services; embedding the test reduces barriers. FS highlight on-site integration reduces drop-off. |
| Testing Environment | On-site activities to support longer waits | PWGT report that additional services such as charging stations, food, and comfortable seating extend acceptable wait time to 90–120 min. FS provide education and health information currently delivered during wait time. |
| Personnel | Involve people with lived experience | PWGT mention that testers with personal lived experience of injection drug use are particularly easy to talk to and understanding. |

Acceptable wait time varied by deployment setting and stakeholder role (Fig. 3). Frontline staff were generally more time-constrained than the people they serve, reflecting their operational perspective. At harm reduction organizations/SSPs and primary care settings, frontline staff endorsement concentrated at 30 minutes or less, while PWGT tolerance extended further, splitting between the 31–60 and 91–120 minute brackets at harm reduction/SSPs and concentrating at 31–60 minutes at primary care. In street outreach and encampment settings, several frontline staff noted that without incentives, patients would not wait longer than 20 minutes. However, interviews with PWGT revealed that offering additional services such as charging stations, food, and comfortable seating could extend acceptable wait times to the 91–120 minute range, suggesting that the operational constraint can be addressed through co-location with existing harm reduction services. In recovery centers and outpatient SUD settings, frontline staff endorsement spanned all brackets from ≤20 to 91–120 minutes, with 31–60 minutes as the largest single segment, while PWGT endorsement split between 31–60 minutes and 91–120 minutes, indicating a gap between tester workflow demands and patient tolerance in this setting. Jails and corrections showed the tightest frontline staff threshold overall, with approximately 20% of frontline staff requiring results within 20 minutes; PWGT tolerance at this setting was markedly higher, with endorsement falling in the 91–120 minute bracket. Frontline staff also noted that the ability to run multiple tests in parallel could meaningfully offset time-to-result constraints. Together, these findings establish that time to result is a setting-dependent specification, driven primarily by frontline staff tolerance rather than patient tolerance; we set ≤30 minutes as the optimal target and ≤60 minutes as the minimal acceptable threshold across primary deployment contexts. These implementation constraints directly shaped the device-level requirements identified by stakeholders and are carried forward into the design specifications presented in Table 3.

**Figure 3.**
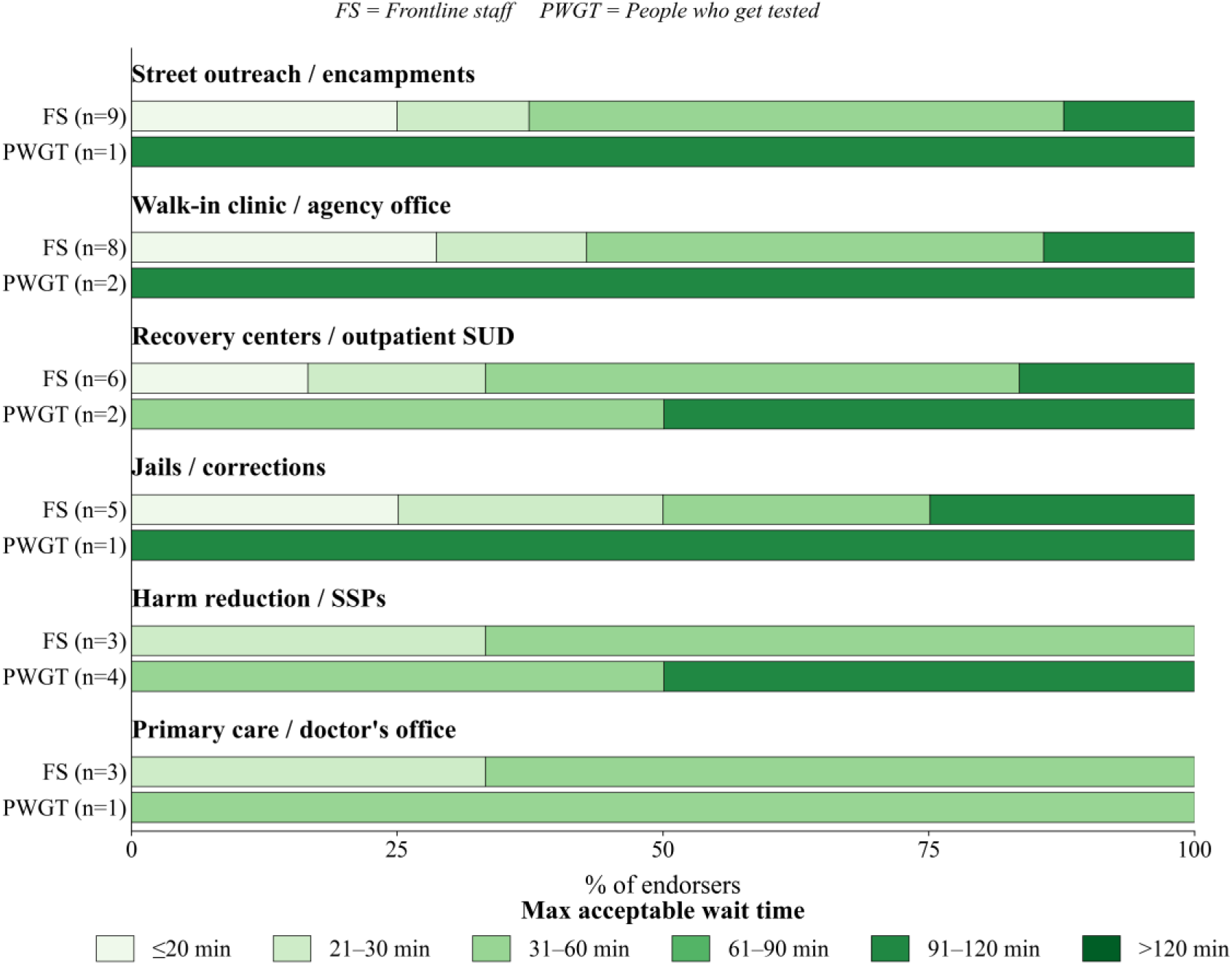
Maximum acceptable wait time for point-of-need HCV RNA test results, by testing setting and stakeholder group. Maximum acceptable wait time for point-of-need HCV RNA test results, reported by frontline staff (FS) and people who get tested (PWGT), stratified by testing setting. Tolerance for wait time varied by setting and stakeholder group. At harm reduction organizations/SSPs and primary care, FS endorsement concentrated at 30 minutes or less. PWGT endorsement varied more by setting: at harm reduction organizations/SSPs, endorsement was split between the 31–60 minute and 91–120 minute brackets; at primary care, endorsement concentrated at 31–60 minutes; at recovery centers/outpatient SUD, endorsement was split between 31–60 minutes and 91–120 minutes; and at street outreach, walk-in clinics, and jails/corrections, a majority or all PWGT endorsement fell in the 91–120 minute bracket.

### Design Considerations

The design considerations discussed in Table 2 were derived from participant perspectives on aspects of their current methods, what works, what does not, things they liked, and challenges currently faced, as well as from the contextual investigation and need assessment. These included turnaround time, sample type and volume, portability, result output, target operator, and ease of use.

**Table 2.**
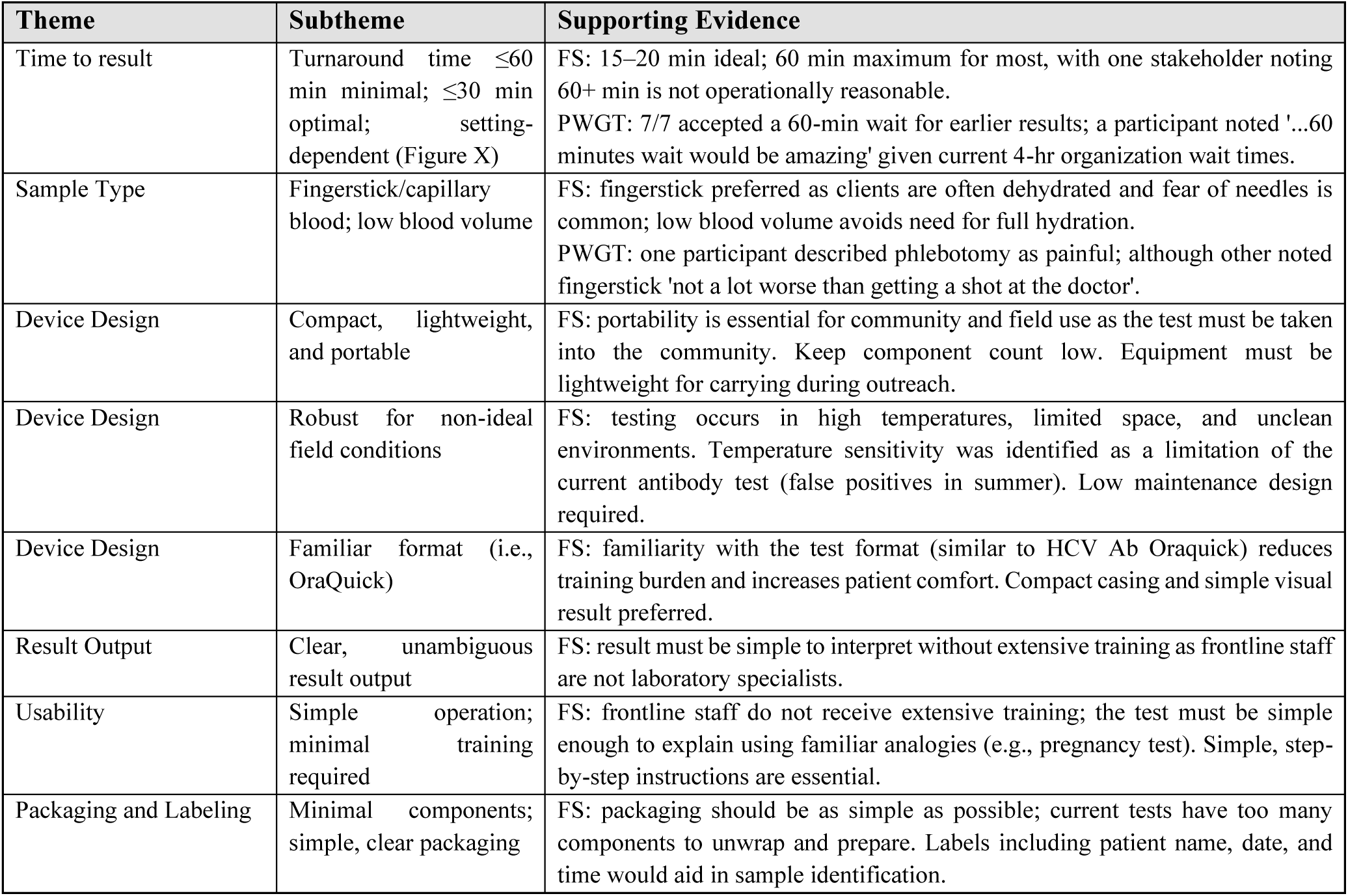
Stakeholder-derived design considerations for a point-of-need HCV RNA test for harm-reduction and community settings in Indiana.

These findings, alongside the implementation considerations described in the previous section, were used to inform the design specifications presented in Table 3.

**Table 3.** Stakeholder-derived design specifications for a point-of-need HCV RNA test for harm-reduction and community settings in Indiana — minimum and optimal targets benchmarked against the FIND Dx HCV TPP (2015).

| User requirement | Design specifications |  | Gap and Finding |
| --- | --- | --- | --- |
|  | Minimum | Optimal |  |
| ANALYTICAL PERFORMANCE |  |  |  |
| Diagnostic sensitivity (vs. NAT gold standard) | 90 – 95% * | ≥ 99% * | TPP range accepted; stakeholders did not modify. 95 – 99% considered programmatically adequate. |
| Diagnostic specificity (vs. NAT gold standard) | > 98% * | > 99% * | TPP minimum confirmed. High specificity critical for a one-step algorithm to avoid false positives leading to unnecessary treatment initiation. |
| Analytical sensitivity (limit of detection) | 1,000 – 3,000 IU/mL * | 200 IU/mL * (TPP); 1,000 – 3,000 IU/mL ** | 1,000 – 3,000 IU/mL detects > 95% of chronically infected individuals. |
| Analytical specificity | No cross-reactivity with HIV-1, HIV-2, HBV, HEV, ART, anti-TB * | Same as minimum * | Stakeholder-confirmed; particularly relevant given high HIV-HCV co-infection rates in PWUD. |
| USABILITY |  |  |  |
| Specimen type | Venous whole blood or plasma * | Fingerstick / capillary blood *, ** | TPP optimum elevated to minimum-acceptable by stakeholders. Dehydration and fear of needles preclude venous draw in field settings (11/11 FS endorsed fingerstick). |
| Sample preparation | ≤ 2 steps * | Integrated / < 2 steps * | TPP minimum confirmed. FS identified excess components as a key usability barrier in community settings. |
| Time to result | < 60 min *,** | < 15 min * (TPP); ≤ 30 min ** (setting-dependent) | Setting-dependent threshold added: ≤ 60 min for SSP / harm reduction (100% endorsement); ≤ 30 min for street outreach and corrections. Multiple concurrent devices and additional services available are recommended for field deployment. |
| Target operator | Health-care worker or laboratory technician with limited training * | Community health worker with minimal training *, ** | Stakeholders specified additional requirements: explainable via familiar analogies (OraQuick); step-by-step printed instructions; no laboratory expertise assumed. |
| Result output / interpretation | Positive / negative readout; simple results screen * | Integrated screen with patient ID, date, location * (TPP); Familiar format (OraQuick) ** | Stakeholders reframed the optimum toward format familiarity rather than an integrated screen. Format familiarity not specified as a design requirement in the TPP. |
| OPERATIONAL & IMPLEMENTATION |  |  |  |
| Instrumentation / portability | Allow separate sample-prep device *,** | Instrument-free * (TPP); Compact, lightweight, handheld ** | TPP portability framed around health-center use. Stakeholders require a handheld design physically carried into the community by frontline staff; mobile deployment not addressed in TPP. |
| **Power requirements** | Rechargeable battery ≥ 8 h *<br>Battery, 2 h per event ** | Battery + solar; circuit protector; ≥ 3 days continuous use * (TPP);<br>Battery, 4 h per event ** | TPP battery duration broadly met. Event-specific framing (2 – 4 h per testing event) not addressed in TPP; no continuous mains power available in field. |
| **Biohazard waste** | No biosafety cabinet; consumables disposable as biosafety waste * | Closed system; recyclable plastics; no open biohazard handling * (TPP); Minimal generation, transportable post-event ** | TPP addresses biosafety broadly but not mobile post-event transport constraints. |
| **Packaging & labelling** | Simple; aliquoted for single use *<br>Patient labels ** | Not specified * (TPP); Minimal components; clear packaging** | Packaging complexity is an active usability barrier. Sample-labelling workflow (patient name, date, time) identified as a workflow opportunity, not addressed as a design specification in the TPP. |
| **Cost per test** | < US \$15 * | < US \$5 * (TPP);<br>≤ US \$42.84 ** (Indiana Medicaid) | TPP price target broadly compatible. Indiana Medicaid reimbursement (\$42.84/test (43)) is the upper bound; state-grant / credit procurement pathway not addressed in TPP. |
| **Testing setting / deployment** | District hospital (Level II) *<br>Organization's office, SSP, harm reduction ** | Community / health center (Level I) * (TPP);<br>Street outreach, jails, libraries, community events ** | TPP lowest setting (community health center) does not encompass mobile, non-facility deployment contexts required for the PWUD population. |
| **Personnel with lived experience** | Not addressed * | Not addressed * (TPP);<br>Testers with lived experience of injection drug use preferred ** | Structural implementation is entirely absent from TPP. PWGT describe testers with lived experience as particularly easy to talk to, which increases engagement and trust. |

\* From FIND Dx high-priority target product profile for HCV diagnosis in decentralized settings (2015). (31)
\*\* From semi-structured stakeholder interviews with Frontline staff (FS), and people who get tested (PWGT), Indiana, 2024.
Gap and Finding describe how stakeholder input confirmed, modified, or extended the FIND TPP specification, or introduced a wholly stakeholder-derived requirement.
NAT, nucleic acid amplification test; PWUD, people who use drugs; SSP, syringe-services program; CHW, community health worker.

### Translating Findings to Design Specifications

The human-centered approach made it clear which testing circumstances and purposes would be most valuable for the diagnosis of the most at-risk populations, the development of a test designed to be deployed at harm reduction and SSP organizations across Indiana.

Together, the design and implementation considerations identified from the interviews were used to create a set of user requirements to inform design specifications. Important criteria that were insufficiently addressed were filled in using public material from the FIND Diagnostic’s target product profile (Table 3). TPP was compared against the design specifications to determine whether each was fulfilled, failed, or was not evaluated in the data presented in this report. Interview data accounted for over 50% of the design specifications as compared to the external reports. This additional information allowed design specifications to be tailored to their intended use case, user, and proposed value far more than the publicly available reports listed above.

## Discussion

Our central finding is that a top-down target product profile, while necessary, is not sufficient to guide the development of a diagnostic intended for deployment in harm-reduction settings. Applying a human-centered design approach, we identified specifications that the FIND Dx HCV TPP either omits, understates, or frames incorrectly for mobile field deployment. Kimani et al. (44) argued that the requirements shaping rapid diagnostic tests routinely overlook the clinicians, community health workers, and patients most critical to their adoption; our work operationalizes that critique by naming the specific requirements a top-down process failed to surface for this population, and by translating them into engineering-actionable design specifications in Table 3. The minimum-versus-optimal framing of the FIND TPP breaks down in three ways when tested against real-world deployment. First, some TPP requirements framed as optimal were found to be the minimum needed for proper implementation. For example, fingerstick capillary blood is listed as an optimal specimen type, but every frontline staff member (11/11) and every person interviewed after receiving testing (7/7) endorsed fingerstick as the minimum acceptable specimen given dehydration and fear-of-needles concerns. Second, several TPP minima are non-negotiable in ways the TPP does not flag, no access to temperature-controlled environments, no continuous power supply, and post-event biohazard transport are field-defining constraints, not preferences to be optimized against. Third, several requirements are absent from the TPP entirely, most notably the value of frontline staff with lived experience as the test operator, patient-labeled sample workflow, and a state-grant or credit-based procurement pathway that matches how these organizations actually acquire supplies. For diagnostic developers, the practical implication is that TPPs should be treated as scaffolding, not a completed requirements set. Thus, a context-specific, stakeholder-derived layer should be developed for the intended deployment environment and integrated into design decisions from the outset.

Time-to-result is typically specified in TPPs and product briefs as a fixed number; our findings suggest it is more accurately modeled as a function of deployment context. Frontline staff working in street-outreach and encampment settings reported that clients would not wait longer than approximately 20 minutes without incentives, whereas people who use harm-reduction services in this setting predominantly endorsed the 91–120 minute bracket, and interviews further indicated that co-locating charging stations, food, and comfortable seating helped sustain this level of tolerance. The practical implication for developers is that a single fast device is not always the right target: in outreach settings, several parallel devices with a longer per-run time may be operationally superior to a single faster device. Time-to-result specifications should therefore be defined by deployment setting, not as a single global threshold.

Participants who use harm-reduction services described frontline staff with lived experience of injection drug use as particularly easy to talk to and instrumental to their engagement with testing. This aligns with prior work by Muncan et al. (27) documenting that stigma from healthcare workers is a primary barrier to healthcare engagement among people who inject drugs in urban US settings, and with intersectionality-lens analyses of hepatitis C point-of-care testing that identify trust in the operator as a determinant of testing uptake (45,46). The FIND TPP frames the target operator only in terms of training level (healthcare worker, CHW). Our findings suggest that the operator role itself is a design specification: a device intended for this population should be designed for training and deployment adhering to current personnel training timelines and processes. This has concrete design consequences, including the use of familiar test-format analogies (OraQuick) in the development of instructions, and avoidance of clinical or scientific terminology.

Existing near-patient HCV RNA platforms, including the FDA-authorized Cepheid Xpert HCV Viral Load Fingerstick assay (18,19) and the Genedrive and Truenat platforms (20–24), have demonstrated analytical performance adequate for decentralized testing but when assessed against the stakeholder-derived specifications developed here, none of these platforms can be implemented by the mobile frontline staff working across different community settings. This clarifies why ‘point-of-care’ as currently instantiated has not translated into improved diagnosis and treatment rates among people who use drugs (33,34) despite a decade of TPP-guided device development. A distinct design target is required to close this gap: point-of-need, defined by mobility and infrastructure independence. The stakeholder-derived specifications in Table 3 provide a first articulation of what that target looks like in practice.

## Conclusion

Stakeholder-derived design requirements for a point-of-need HCV RNA test in harm-reduction settings extend and refine the FIND Dx TPP by adding specifications that a top-down process could not surface: fingerstick as a minimum rather than an optimal specification, time-to-result as a setting-dependent specification, involvement of personnel with lived experience as a design requirement, and procurement and mobile-transport constraints that shape what the device must be. TPPs remain a valuable scaffold for early-stage development, but they are not a substitute for direct engagement with the frontline staff and community members who determine whether a device is ultimately adopted. For diagnostic developers working with marginalized populations, we recommend that a context-specific, stakeholder-derived requirements set be treated as a required layer above the TPP and integrated into the design process from the outset, rather than validated against user needs only after a prototype exists.

## Limitations

This study was conducted in a single US state (Indiana) with 11 frontline staff and 7 individuals who use harm-reduction services. The people-who-get-tested sample identified as White/Caucasian or did not report race, and the frontline sample was recruited through snowball sampling seeded within the research group’s existing network. Findings may differ in large-metropolitan sites with denser harm-reduction infrastructure, and in populations with different racial and ethnic composition where the intersection of drug use, incarceration history, and healthcare access differs from the Indiana context (45–47).

This study should be interpreted as formative. RQA, by design, uses structured coding matrices mapped to predefined domains of interest instead of the iterative, sample-expanding theoretical sampling that underlies data saturation in grounded theory (39). We assessed sample adequacy using prior published studies that use the same methodology or a similar approach and materials (48–50), and paired-coder consensus review across both matrices supported the depth and consistency of coding for the domains examined; the study was not, however, powered to detect effects across subgroups. Together, these parameters support the adequacy of an 11-participant and 7-participant sample for generating an initial set of stakeholder-derived design requirements. We present these findings as a formative design-requirements baseline, to be tested and refined as we iterate through the HCD process.

Because the device does not yet exist, acceptability and design responses reflect stated preferences elicited against a described concept rather than observed use of a working prototype; observational studies with a functional prototype in field settings will be conducted in future work to confirm and refine these requirements. This analysis is grounded in the inspiration and ideation phases of HCD; the implementation phase, including iterative prototype testing with the same stakeholder groups, remains future work.

Wait-time tolerance findings should be interpreted with caution given the small number of endorsers within several setting-by-stakeholder-group categories (e.g., n=1 to n=3 for some FS and PWGT subgroups), meaning a single participant’s response can shift the reported percentage substantially. These values were derived from qualitative coding of participants’ stated experiences and preferences during semi-structured interviews, rather than from a validated quantitative instrument designed to measure wait-time tolerance directly. As such, the percentages presented in Figure 3 should be read as indicative of directional patterns across settings rather than as precise population estimates, and findings for the smallest subgroups in particular warrant confirmation in a larger sample before being used to inform specific design thresholds.

## Supporting information

COREC Checklist

Interview Guide FS

Interview Guide PWUD

## Data Availability

All de-identified data produced in the present study will be available upon reasonable request to the authors

## Abbreviations

CHW: community health worker;
FDA: U.S. Food and Drug Administration;
HCD: human-centered design;
HCV: hepatitis C virus;
NAT: nucleic acid amplification test;
PARRQA: Planning for and Assessing Rigor in Rapid Qualitative Analysis;
POC: point-of-care (existing devices/landscape);
PON: point-of-need (target of the device under development);
PWGT: people who get tested;
PWUD: people who use drugs;
RQA: rapid qualitative analysis;
SSP: syringe-services programs;
TPP: target product profile.

## Author Contributions

SMR determined the methodological approach, conducted interviews, served as primary analyst for both stakeholder groups (paired with JK for one group and AC for the other), and led manuscript writing. KK conducted interviews, served as the third reviewer across both groups, and contributed to writing and figure development. JK served as co-primary analyst for one stakeholder group and contributed to figure development. AC served as co-primary analyst for the other stakeholder group and contributed to writing. LB completed RQA methodology training, reviewed RQA analytic materials and the interview guide prior to deployment and contributed writing feedback. JCL (BME PI) and NMR (PUBH PI) supervised all stages of the project, obtained IRB approval and led subsequent IRB revisions, provided methodological guidance throughout study design and analysis, contributed to manuscript framing, and reviewed the manuscript.

## Acknowledgement

Funding for this study included support by the National Institutes of Health National Institute on Drug Abuse award # DP2DA051910.

