## Supplementary material for "Beyond the Target Product Profile: Design Requirements for Point-of-Need HCV RNA Testing in Harm Reduction Settings": Interview Guide FS

### DP2 Questionnaire for <u>stakeholders</u> who do HCV testing.

Principal Investigator: Dr. Jacqueline Linnes
Primary Contact: Samantha Mata Robles

**Introductory Script**

This interview is to learn your thoughts and experiences related to Hepatitis C screening. We want to hear what you think about current testing methods and potential alternatives. There are no right or wrong answers. We may ask you questions that you don't know the answer to and that's okay. Your opinions are very important to us. They will help us improve Hepatitis C screening.

**Questions**

1. Target Clientele
    a. Who does your organization serve?
    b. How many people do you serve?
    c. What kind of recruitment/outreach activities are done?
    d. How many resources are allocated to that? (time, people, budget)
    e. How is success measured?
        i. Are there goals to be met in terms of tests done, events, etc.?
    f. What are some barriers with connecting and engaging the community?
2. Testing
    a. What are the current HepC testing guidelines that the organization you work for follows? (CDC, FDA, Indiana Health Decisions)
    b. How did you become a tester?
        i. What does the training process involve?
    c. Which tests are currently used?
    d. Can you walk me through the process when a patient comes to get tested? as specific as possible please.
        i. Testing steps?
        ii. How long does the process last for a single patient?
        iii. How long are your overall testing visits/events?
        iv. What are some things you **like** about the current test and protocol you use?
            1. (Ease of use, time, testing process [before during and after])
        v. What are things you **dislike** about the current test and protocol you use?
            1. (Ease to use, time, testing process [before during and after])
        vi. What are some challenges with using the tests with your target audience?
        vii. What is the current wait time between starting a test and getting the results?
            1. What do you do in the meantime?
        viii. What changes would you want to make to the process?
        ix. What are some considerations when testing amongst PWUD or at-risk populations?
            1. Do they need to be incentivized?
            2. Do you have to go to them, or do they go to you? (walk in hours/events)

e. Walk me through what it's like after someone has been tested?
    i. What services are they offered?
        1. What is the process like and along which parts can we lose the patients?
            a. Why?
    ii. How do you encourage patients to stay involved?
    iii. How do you keep track of the patients and their health status?
    iv. Is there anything done to address other challenges that they might face like housing, access to food, education, overall health?

3. Needs
    a. Do you know who determines the tests you use?
    b. Do you know how much current tests you use cost?
        i. Is price per test a factor that influences the decision of which one to use?
    c. How does the procurement for the tests work?
    d. What does the test kit need to include?
    e. How would an ideal test look for you? How would you improve on the current test?
    f. What strains of HepC do you test for?

4. Our Test

Our team of engineers is currently developing a rapid test that would directly detect the virus (like lab tests do) instead of detecting antibodies (like what current rapid tests do).

a. What are your initial thoughts on that?
b. Would this be something your organization and the population could benefit from?
c. What would the maximum feasible cost of the test be?
d. If the results were positive, how would you integrate them to care?
e. If the test could detect acute HCV at least a month earlier than antibody tests but takes about 60 minutes to run, do you think this trade-off is worthwhile?
    i. Do you think it be beneficial to directly detect the virus instead of the antibodies at your testing stage?
    ii. Do you think there is even a need to be able to detect HCV earlier for PWUD/other at-risk populations?
    iii. What would be the maximum time this test could take for you to feasibly implement it?
f. Is there anything else you'd like for test developers to know or keep in mind as they work to design or implement this new test?

**Concluding Script**

Those are all the questions I have for you today. Thank you again for your time in this interview. Please don't hesitate to follow up with any questions or concerns! [*proceed to gift card process*]
