## Supplementary material for "Beyond the Target Product Profile: Design Requirements for Point-of-Need HCV RNA Testing in Harm Reduction Settings": Interview Guide PWUD

### DP2 Questionnaire for <u>people who get tested</u>.

Principal Investigator: Dr. Jacqueline Linnes
Primary Contact: Samantha Mata Robles

#### Consent form

Before beginning the study, I'll read to you the consent form. If you have any questions let me know and we can go through them. After, I'll ask you some questions about what the study entails.

*(reads consent form)*

#### Capacity of Consent Assessment

*After reviewing the consent form, ask the following questions:*

1. If you decide to be in this study, what are some things you will be asked to do?
2. What are some risks of being part of the study?
3. What are some benefits of being part of the study?
4. Do you have to participate in this study?
5. What will happen if you decide not to be in the study?

*<u>If consent capacity is not reached</u>, then direct to Sam or "Thank you for your interest in this study. Unfortunately, you cannot participate at this time. Please get back in touch with Sam."*

*<u>If consent capacity is reached:</u> Continue with study to introductory statement*

#### Introductory statement

This interview is to learn your thoughts and experiences related to Hepatitis C. We want to understand what makes it difficult or easy for you to get tested for Hepatitis C. We want to hear what you think about current testing methods and potential alternatives. There are no right or wrong answers, we may ask you questions that you don't know the answer to and that's okay. Your opinions are very important to us as they will help us improve Hepatitis C testing.

##### Background

Thank you for agreeing to answer some of our questions. We will start with a few on your background.

1. Gender:
   ☐ Male ☐ Female ☐ I choose not to disclose.

2. Age group:
   ☐ 18-24yrs ☐ 25-34 yrs ☐ 35-44 yrs ☐ 45-54 yrs ☐ 55-64 yrs
   ☐ 65-74 yrs ☐ >75 yrs ☐ I choose not to answer.

3. Education: What level of education have you achieved?
   - Less than 12 years of schooling.

- High school graduate
- Vocational education in ___________________
- Bachelor's degree
- Master's
- Doctorate
- Other special training or certification ____________________

4. What is your ethnicity?
   - African American
   - White non-Hispanic
   - Hispanic
   - Native American
   - Asian Pacific Islander
   - Other ____

5. Are you currently homeless? (yes/no)

6. Do you currently have health insurance? (yes/no)

7. Are you currently employed? (yes/no)
   a. Annual Income?

8. Injection Drug Risk: Have you ever injected drugs? Yes/no/prefer not to answer.
   a. If yes, in the past month?
      i. How many times did you inject drugs in the past month?
      ii. Of the times that you injected drugs, what proportion of those times did you share a needle or syringe? (never, rarely, sometimes, half, most, always/almost always)

9. Sexual Risk: How many sexual partners have you had in the past year?
   a. Do you know your partner(s) HepC status? (yes/no)
   b. Have you ever had an HepC-positive sex partner? (yes/no)
   c. What proportion of the time did you or your partner(s) use a condom while engaging in sexual activity? (never, rarely, sometimes, half, most, always/almost always)
   d. Have you had unprotected sex in the past 6 months? (yes/no)

#### Questions

1. Patient's beliefs:
   a. Have you heard of Hepatitis C?
      *i.* **If 'no'*:*** *According to the CDC, HepC is a liver infection caused by the hepatitis C virus (HCV). Hepatitis C is spread through contact with blood from an infected person. People at increased risk for hepatitis C are: People who use injection drugs or did so in the past, even those who injected only once many years ago; People with HIV infection; People with certain medical conditions; People who have received transfusions or organ transplants before 1987; Health care,*

*emergency medical, and public safety personnel who have been exposed to the blood of someone who has hepatitis C (through needle sticks, sharps, or mucosal exposures); and Children born to mothers who have hepatitis C.*

1. Based on the CDC definition, do you have any questions or concerns about HepC?

ii. **If 'yes'**:

1. How did you learn about HepC?
2. Do you think you might be at risk for HepC?

*According to the CDC, HepC is a liver infection caused by the hepatitis C virus (HCV). Hepatitis C is spread through contact with blood from an infected person. People at increased risk for hepatitis C are:People who use injection drugs or did so in the past, even those who injected only once many years ago, People with HIV infection, People with certain medical conditions, People who have received transfusions or organ transplants before 1987, Health care, emergency medical, and public safety personnel who have been exposed to the blood of someone who has hepatitis C (through needle sticks, sharps, or mucosal exposures), and Children born to mothers who have hepatitis C.*

3. Based on your previous knowledge and the CDC definition, do you have any questions or concerns about HepC?

b. Have any friends, family, or yourself had any experiences with HepC? (please describe if you are comfortable)
   i. Do you perceive it to be a serious disease?

2. Testing
   a. Have you ever gotten tested for HepC?
      i. Why or why not?
   b. When was the last time you got tested?
      i. Where did you get tested?
      ii. Walk me through the process.
         1. What do you think about the test? In terms of wait times, convenience, pain, etc.
   c. Do you find it easy or difficult to access testing?
      i. How so?
      ii. What kind of things make getting tested challenging for you?
      iii. What would make it easier for you to get tested?
   d. What would the ideal test and process look like for you?
      i. Where would you want to get tested?
      ii. Who would you want to perform the test?
   e. The CDC recommends that people who are at-risk for HepC get tested every 3 months.
      i. **If 'no' to risk question:** what do you think might help or encourage high risk populations to get tested every 3 months?
      ii. **If 'yes' to risk question:** What would make you more likely to get tested every 3 months?

3. Integration to care – starting HepC treatment:
   a. Have you, a friend, or family member ever received HepC treatment?
      **i. If yes:**

1. How did they go from getting a positive test to getting care?
            2. What were some challenges faced?
            3. What things made it easier?
            4. How was the relationship between the provider and the patient?
    b. If you were to have or have HepC, do you think you have the ability to pursue HCV care and treatment if you choose to do so?
        i. What would make you more likely to get treatment if needed?

4. Our test

Our team of engineers is currently developing a rapid test that would directly detect the virus (like lab tests do) instead of detecting antibodies (like what current rapid tests do).

a. If the test could detect HCV at least a month earlier than antibody tests but takes about 60 minutes to run, do you think this trade-off is worthwhile?
b. What would be the maximum time this test could take for you to want to take it?
c. Do you have any additional thoughts on this test?
d. Is there anything else you'd like for test developers to know or keep in mind as they work to design or implement this new test?

**Concluding Script**

Those are all the questions I have for you today. Thank you again for your time in this interview. Please don't hesitate to follow up with any questions or concerns! [*proceed to gift card process*]
